# Serological Response to Three, Four and Five Doses of SARS-CoV-2 Vaccine in Kidney Transplant Recipients

**DOI:** 10.1101/2022.03.23.22270017

**Authors:** Bilgin Osmanodja, Simon Ronicke, Klemens Budde, Annika Jens, Charlotte Hammett, Nadine Koch, Evelyn Seelow, Johannes Waiser, Bianca Zukunft, Friederike Bachmann, Mira Choi, Ulrike Weber, Bettina Eberspächer, Jörg Hofmann, Fritz Grunow, Michael Mikhailov, Lutz Liefeldt, Kai-Uwe Eckardt, Fabian Halleck, Eva Schrezenmeier

**Author notes:** **Corresponding Author:** Bilgin Osmanodja, MD, Department of Nephrology and Medical Intensive Care, Charité – Universitätsmedizin Berlin, Charitéplatz 1, Berlin, 10117 Germany. these authors contributed equally (Bilgin Osmanodja, Simon Ronicke, Fabian Halleck, Eva Schrezenmeier).

## Abstract

Mortality from COVID-19 among kidney transplant recipients (KTR) is high, and their response to three vaccinations against SARS-CoV-2 is strongly impaired.

We retrospectively analyzed serological response of up to five doses of SARS-CoV-2 vaccine in KTR from December 27, 2020, until December 31, 2021. Particularly, the influence of different dose adjustment regimens for mycophenolic acid (MPA) on serological response to fourth vaccination was analyzed.

In total, 4.277 vaccinations against SARS-CoV-2 in 1.478 patients were analyzed. Serological response was 19.5% after 1.203 basic immunizations, and increased to 29.4%, 55.6%, and 57.5% in response to 603 third, 250 fourth and 40 fifth vaccinations, resulting in a cumulative response rate of 88.7%.

In patients with calcineurin inhibitor and MPA maintenance immunosuppression, pausing MPA and adding 5 mg prednisolone equivalent before the fourth vaccination increased serological response rate to 75% in comparison to no dose adjustment (52%) or dose reduction (46%). Belatacept-treated patients had a response rate of 8.7% (4/46) after three vaccinations and 12.5% (3/25) after four vaccinations.

Except for belatacept-treated patients, repeated SARS-CoV-2 vaccination of up to five times effectively induces serological response in kidney transplant recipients. It can be enhanced by pausing MPA at the time of vaccination.

## Introduction

At the beginning of 2021 the successful vaccine campaign against severe acute respiratory syndrome coronavirus 2 (SARS-CoV-2) started, offering protection against hospitalization and death from coronavirus disease 2019 (COVID-19) for most patient groups irrespective of emerging variants.^1^ Despite this success in the general population, solid organ transplant (SOT) recipients had a poor response to vaccination and a limited benefit from the initially recommended two vaccinations.^2^ At the same time the mortality of SOT recipients acquiring COVID-19 is unacceptably high, with rates up to 20% reported in registries.^3,4^ Recent data from the United Kingdom collected between September 2020 and August 2021 show an unadjusted COVID-19 case fatality rate of 9.8% in SOT. The vaccination with two doses did not prevent infections in SOT recipients and increased 28-day survival in COVID-19 infected SOTs only marginally from 88.8% to 91.8%. This is equivalent to a 20% reduction in risk of death in vaccinated SOT recipients, as compared to a 68-fold reduction of death in the general population.^5^

Early in 2021, it was recognized that SOT and especially kidney transplant recipients (KTR) show a diminished serological response compared to healthy individuals and hemodialysis patients.^6-8^ Subsequent studies revealed that the level of antibodies correlates with protection from disease,^9,10^ arguing for serological response controls in SOT recipients.

After three vaccinations, which were early recommended for SOT recipients and later for the general population, serological vaccine response was inadequately low in at least 40%.^11^ T cell response over time only changes at a functional level, never reaching the level of healthy individuals.^12,13^ In summary the poor T cell response in combination with an impaired humoral response offered only a limited protection from infection and a severe course of COVID-19.^14^

The question therefore arises about optimal management of non-responding patients and in particular whether repeated vaccinations increase serological response rates. In the current study, we provide the first systematic investigation analyzing the serological response to up to five repeated vaccinations against SARS-CoV-2 in non-responding KTR. In particular, we report the response rates of KTR after basic immunization, three, four and five vaccinations, and the predictors of serological response after three and four vaccinations as well as the effects of different immunosuppressive reduction regimes on the serological response.

## Methods

### Study population

At our institution, basic immunization against SARS-CoV-2 was performed with two doses of one of the following vaccines in different combinations - BNT162b2 (Comirnaty, BioNTech/Pfizer), mRNA-1273 (Spikevax, Moderna Biotech), ChAdOx1-S (AZD1222, AstraZeneca) or Ad26.COV2.S (Johnson & Johnson, Janssen). Sustained non-responders received up to five doses of SARS-CoV-2 vaccines. A detailed institutional protocol is provided in **Item S1** and **Figure S1**. All patients provided written and informed consent into off-label use for vaccine doses four and five. For the current analysis we included adult kidney transplant recipients, who received SARS-CoV-2 vaccinations from December 27, 2020 until December 31, 2021. Serological response to vaccination, demographic data, transplantation data, medication, as well as routine laboratory data were analyzed retrospectively. The ethics committee of Charité – Universitätsmedizin Berlin approved this study (EA1/030/22).

### Outcome

The primary outcome was serological response to immunization, defined as the maximum serological response after a minimum of 14 days after each immunization, i.e. after basic immunization, three, four and five doses of SARS-CoV-2 vaccines.

We used an anti-SARS-CoV-2 enzyme-linked immunosorbent assays (ELISA) for the detection of IgG antibodies against the S1 domain of the SARS-CoV-2 spike (S) protein in serum according to the instructions of the manufacturer (Anti-SARS-CoV-2-ELISA (IgG), EUROIMMUN Medizinische Labordiagnostika AG, Lübeck, Germany).^15,16^ Processing and measurement were done using the fully automated „Immunomat” (Institut Virion\Serion GmbH, Würzburg, Germany). Results were determined by comparing the obtained signals of the patient samples with the previously obtained cut-off value of the calibrator. As suggested by the manufacturer, samples with a cut-off index ≥ 1.1 were considered to be positive. Alternatively, the electrochemiluminescence immunoassay (ECLIA, Elecsys, Anti-SARS-CoV-2, Roche Diagnostics GmbH, Mannheim, Germany) was used either alone or in parallel detecting human immunoglobulins, including IgG, IgA and IgM against the spike receptor binding (RBD) domain protein. Results were determined by comparing the obtained signals of the patient samples with the previously obtained cut-off value of the calibrator. As suggested by the manufacturer, samples with a cut-off index ≥ 264 U/ml were considered to be positive as recommended by Caillard et al.^17^

Any non-negative titer below the cut-off in each test was defined as low-positive. Accordingly, response to immunization was categorized as sufficient serological response (responders) in case of positive SARS-CoV-2 antibody titer or as insufficient serological response (non-responders) in case of negative or low-positive SARS-CoV-2 antibody titer. The serological response rate was calculated as the rate of responders after each basic immunization, three, four and five doses.

In order to exclude patients with a history of COVID-19, we simultaneously measured antibodies against the nucleocapsid (N) protein with an electrochemiluminescence immunoassay (ECLIA, Elecsys Anti-SARS-CoV-2, Roche Diagnostics GmbH). As before, results were determined by comparing the obtained signals of the patient samples with the previously obtained cut-off value of the calibrator. As suggested by the manufacturer, samples with a cut-off index ≥ 1.0 were considered to be positive.

We included immunization from all adult KTR at our institution within the study period. Immunizations were excluded from analysis when they occurred before transplantation, when no medication data were available, or in case of ineligible serological data (see **Table 1**). The latter occurred if patients received additional vaccination doses without assessment of SARS-CoV-2 IgG titer before and after, mostly when performed outside the transplant center.

**Table 1:**
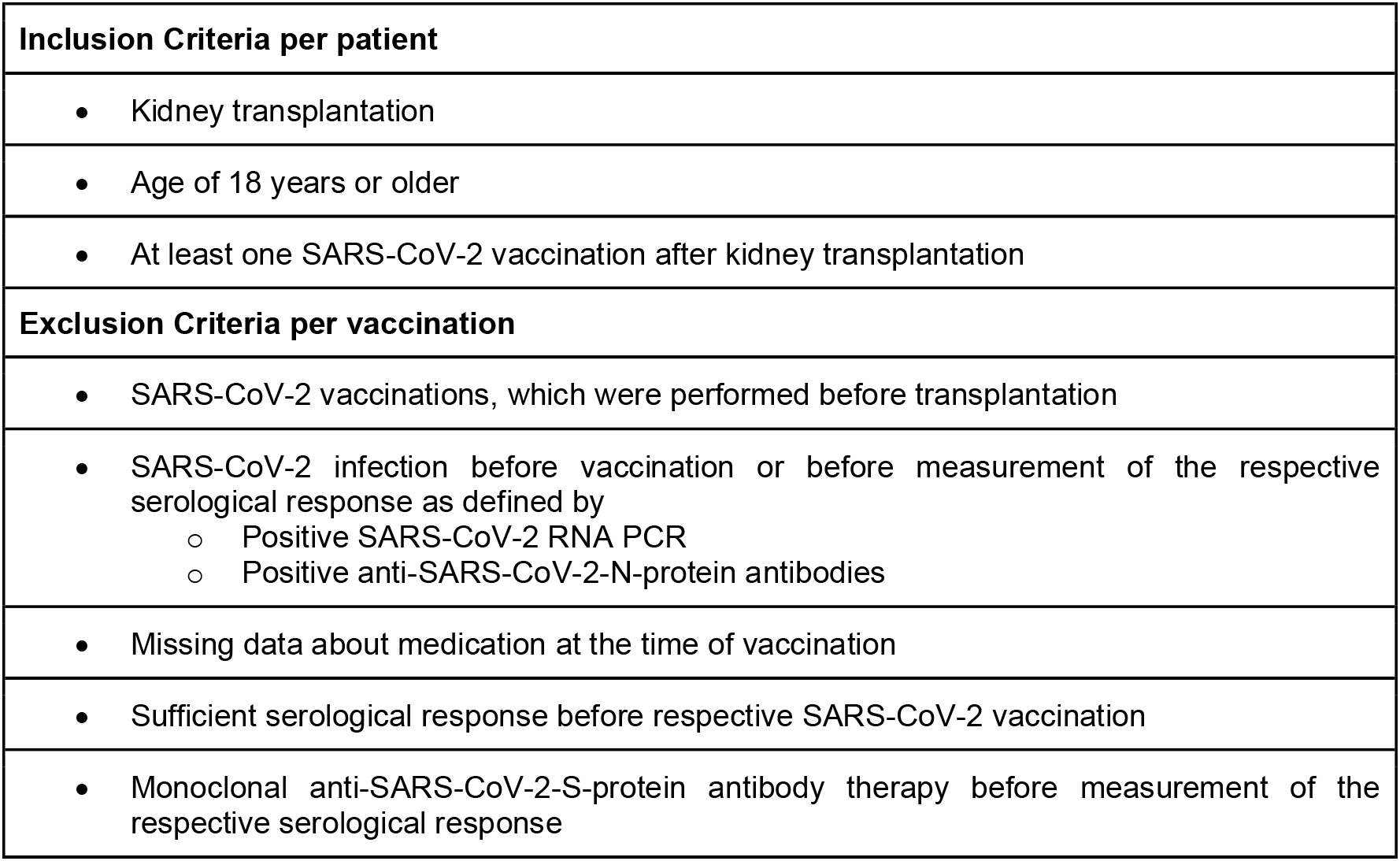
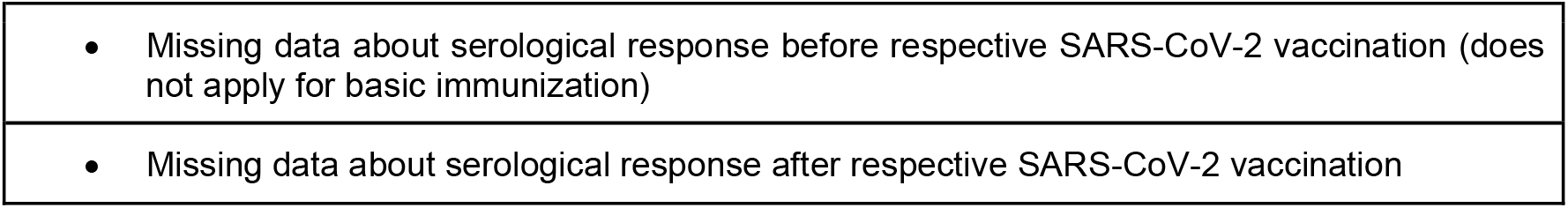
Inclusion and exclusion criteria. Since main data analysis was performed at the vaccination level, all SARS-CoV-2 vaccinations for patients meeting the inclusion criteria were included, while vaccinations were excluded based on the exclusion criteria.

Cumulative serological response was calculated using Kaplan-Meier method with number of vaccinations as time variable and first positive serological response as event of interest. Patients, who remained sustained non-responders after their last vaccination, were treated as censored at this point.^18^

### Multivariable analysis of predictors of serological response

The influence of 19 candidate variables on the primary outcome after the third and fourth vaccination dose was examined in two separate multivariable analyses using logistic regression. Candidate variables included basic patient demographics, transplantation data, vaccination characteristics, latest immunosuppressive medication and routine laboratory parameters (detailed variable definition provided in **Table S1**). Immunizations with missing candidate variable data were excluded from multivariable analysis. No imputation methods were used.

### Comparison of mycophenolic acid (MPA) dose adjustment regimens

Due to the important role of MPA with regard to the response to immunization, different approaches for MPA dose adjustment before fourth SARS-CoV-2 immunization were followed at our institution according to the patients’ individual risk factors such as previous rejection episodes, anti-HLA antibodies, previous response to SARS-CoV-2 immunization, and based on the physicians’ and patients’ shared-decision making. In CNI-treated patients, MPA was reduced or paused from one week before immunization until four weeks after immunization. Steroids were maintained at 5 mg prednisolone equivalent. In case of steroid-free treatment 5 mg prednisolone equivalent was added for the time of MPA reduction, which was discontinued after restart of MPA.

To examine MPA related effects, patients on CNI-based immunosuppression receiving a fourth dose of a SARS-CoV-2 vaccine were assigned to three groups according to their change in MPA dose in relation to the MPA dose before their third SARS-CoV-2 vaccination: (1) steady MPA dose, (2) reduced MPA dose and (3) paused MPA. Serological response rates were compared between groups using Mann–Whitney U test. Continuous variables between groups were compared using t-test.

### Serological response in patients with CNI and belatacept maintenance immunosuppression

The serological response rate in patients on CNI-based as well as belatacept-based immunosuppression was calculated separately. Additionally, cumulative serological response rate was described as stated above. For patients with belatacept-based immunosuppression, no cumulative serological response rate is shown after 5 vaccinations, due to the low patient count in this group. Serological responders receiving belatacept-based immunosuppression were further analyzed on the patient-level.

Statistical analysis was performed using R studio v.1.2.5042 and R version 4.0.2 (2020-06-22).

## Results

### Serological response to immunization against SARS-CoV-2

A total of 4.277 vaccinations against SARS-CoV-2 in 1.478 patients were evaluated. The distribution of included and excluded immunizations, the reasons for exclusion and the resulting serological response is shown in the patient flow diagram (**Figure 1**). Demographic, clinical and vaccination data for patients receiving three, four and five doses of SARS-CoV-2 vaccines are summarized in **Table 2**.

**Figure 1:**
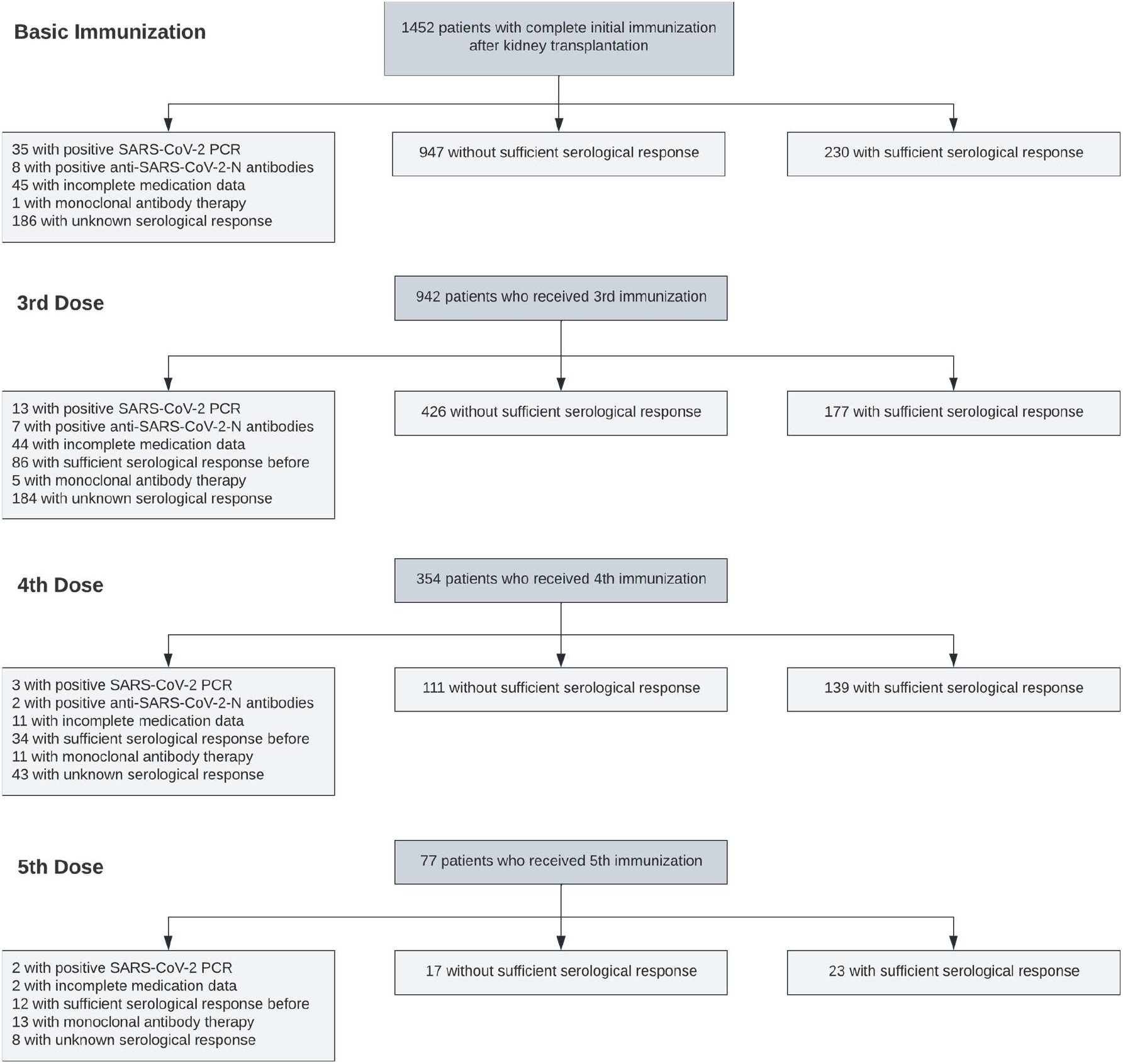
Patient flow diagram showing the number of patients included into each analysis and the number of excluded patients and the respective reasons for exclusion.

**Table 2:** Baseline characteristics of patients who received three, four and five doses of SARS-CoV-2 vaccines. Continuous variables are presented as mean (+/- standard deviation) unless stated otherwise. IQR – interquartile range. BMI – body mass index. IS – immunosuppression. mTORi – mammalian target of rapamycin inhibitor. eGFR – estimated glomerular filtration rate.

| **Vaccination Number** | **3** | **4** | **5** |
| --- | --- | --- | --- |
| Total Patients | 603 | 250 | 40 |
| **Demographics and Comorbidities** |  |  |  |
| Female / male patients | 38% / 62% | 33% / 67% | 45% / 55% |
| Median age in years (IQR) | 59 (48 - 68) | 61 (51 - 70) | 63 (56 - 72) |
| BMI in kg/m<sup>2</sup> | 25.4 +/- 4.7 | 25.0 +/- 4.4 | 24.76 +/- 4.77 |
| Diabetes | 21.7% | 21.8% | 18.0% |
| **Transplantation** |  |  |  |
| Median transplant age in years (IQR) | 8.2 (3.1 - 13.5) | 7.7 (3.0 - 12.7) | 7.2 (2.2 - 11.5) |
| Tacrolimus-based IS | 73.6% | 73.6% | 77.5% |
| Ciclosporin-based IS | 16.8% | 16.4% | 12.5% |
| Belatacept-based IS | 7.6% | 10% | 12.5% |
| Patients with MPA | 93.7% | 50.4% | 42.5% |
| Patients with mTORi | 1.0% | - | - |
| Patients with Azathioprine | 0.8% | - | - |
| Patients with more than 2 immunosuppressive drugs | 60.7% | 39.6% | 32.5% |

| **Laboratory Values** |  |  |  |
| --- | --- | --- | --- |
| Baseline eGFR in ml/min/1.73m<sup>2</sup> | 51.2 +/- 20.0 | 47.5 +/- 19.6 | 43.35 +/- 18.68 |
| Urine albumin creatinine ratio in g/g | 0.14 +/- 0.45 | 0.19 +/- 0.62 | 0.44 +/- 1.45 |
| Hemoglobin in g/dl | 12.68 +/-1.65 | 12.56 +/- 1.52 | 12.73 +/- 1.69 |
| Leukocyte count in /nl | 7.33 +/- 2.38 | 7.41 +/- 2.53 | 7.86 +/- 2.44 |
| **Vaccination** |  |  |  |
| Baseline SARS-CoV-2 IgG low positive | 3.5% | 11.6% | 45% |
| mRNA Vaccination | 72.8% | 86.8% | 100% |
| Median time since previous vaccination in days (IQR) | 71 (53-102) | 64 (55 - 84) | 62 (46 - 70) |

The vaccination-specific rate of serological response to 1.203 basic immunizations that met the inclusion/exclusion criteria was 19.5%. The rate increased to 29.4%, 55.6%, and 57.5% in response to 603 third, 250 fourth and 40 fifth vaccinations against SARS-CoV-2, respectively (**Figure 2A**). Correspondingly, cumulative serological response increased from 19.1% after two vaccinations to 42.0% after three, 74.2% after four, and 88.7% after five vaccinations (**Figure 2B**). No serious adverse events were found in patients receiving fourth or fifth vaccination dose.

**Figure 2.**
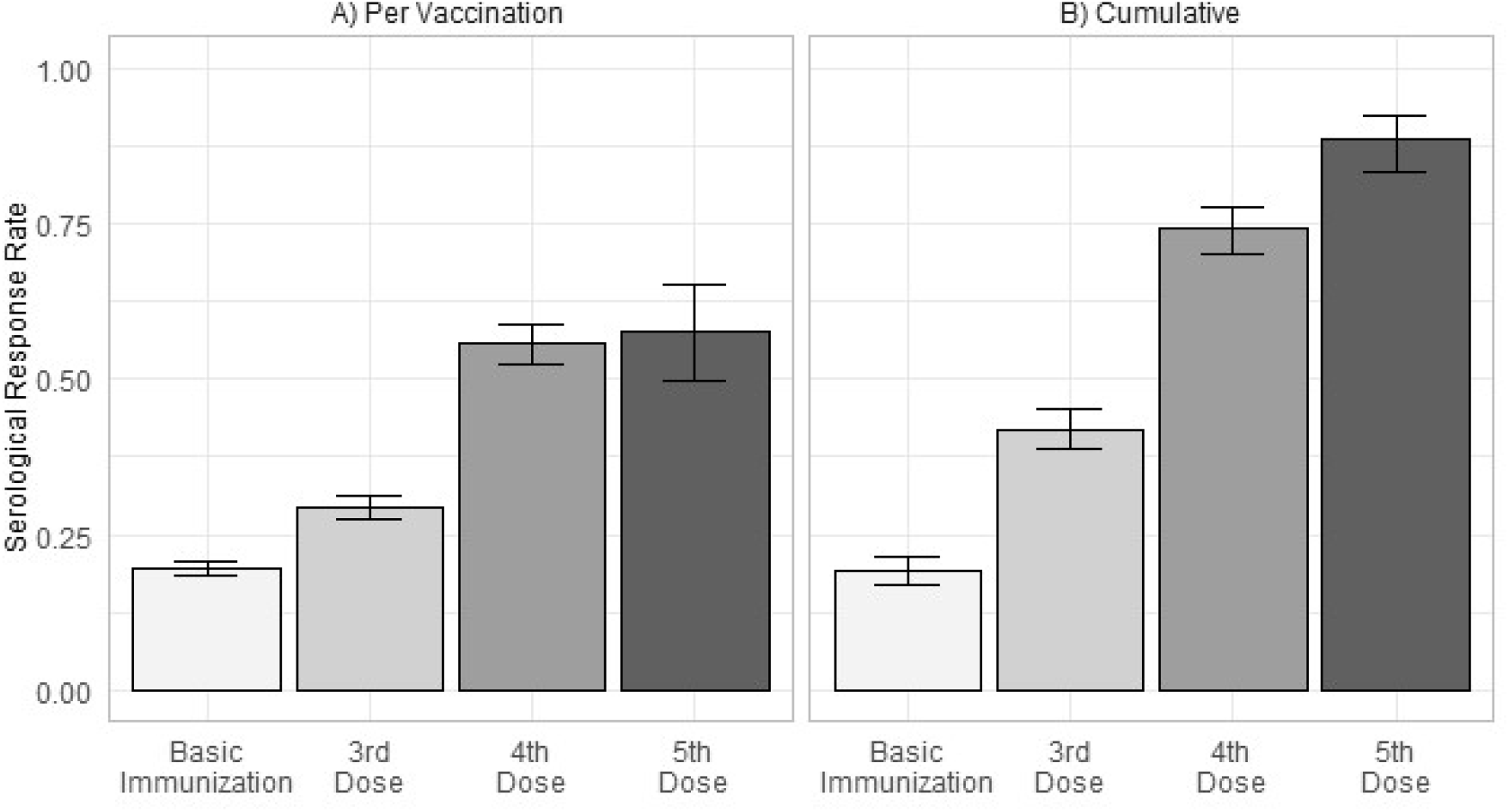
**A)** Serological response rate (+/- standard deviation) per vaccination after basic immunization, three, four and five doses of SARS-CoV-2 vaccines in kidney transplant recipients without sufficient serological response before the latest vaccination. **B)** Cumulative serological response rate (+/- 95% confidence interval) after up to five doses of SARS-CoV-2 vaccines in all kidney transplant recipients with at least one vaccination meeting the inclusion and exclusion criteria.

### Predictors of serological response to immunization against SARS-CoV-2

We performed multivariable analysis using logistic regression for 574 patients with third vaccination and 226 patients with fourth vaccination separately to identify factors that influence serological response. We found any previous low positive anti-SARS-CoV-2-S-protein IgG titer, younger age, higher BMI, higher transplant age, higher estimated glomerular filtration rate (eGFR), and higher hemoglobin levels to be associated with improved serological response after three doses of SARS-CoV-2 vaccine. Younger age, higher transplant age and mRNA-based vaccination were associated with improved serological response after four doses of SARS-CoV-2 vaccine (**Table 3** and **4**).

**Table 3:** Predictors of serological response after three doses of SARS-CoV-2 vaccines identified in multivariable analysis.

| Variable | Odds Ratio (95% CI) | P value |
| --- | --- | --- |
| **Low positive anti-SARS-CoV-2-S-protein IgG before vaccination** | **30.5 (7.31 - 224)** | **<0.001** |
| Female sex | 0.88 (0.54 - 1.40) | 0.59 |
| **Age** | **0.97 (0.96 - 0.99)** | **0.002** |
| **BMI** | **1.06 (1.01 - 1.11)** | **0.019** |
| mRNA vaccine | 0.75 (0.46 - 1.22) | 0.24 |
| **Transplant age** | **1.05 (1.02 - 1.09)** | **<0.001** |
| Diabetes | 1.15 (0.68 - 1.91) | 0.60 |
| CNI treatment | 0.74 (0.15 - 4.22) | 0.71 |
| Steroid treatment | 0.97 (0.32 - 3.07) | 0.96 |
| **Belatacept** | **0.12 (0.02 - 0.85)** | **0.04** |
| MPA | 1.01 (0.24 - 4.42) | 0.99 |
| **MPA dose in MMF equivalent in g** | **0.26 (0.16 - 0.44)** | **<0.001** |
| mTORi | 4.78 (0.49 - 61.0) | 0.19 |
| Azathioprine | 0.28 (0.02 - 3.47) | 0.34 |
| More than 2 immunosuppressive drugs | 0.63 (0.19 - 1.92) | 0.42 |
| **eGFR in ml/min/1.73m<sup>2</sup>** | **1.03 (1.01 - 1.04)** | **<0.001** |
| Leukocyte count | 0.96 (0.87 - 1.06) | 0.41 |
| **Hemoglobin** | **1.28 (1.11 - 1.49)** | **<0.001** |
| Albumin creatinine ratio in g/g | 0.71 (0.27 - 1.32) | 0.39 |

**Table 4:** Predictors of serological response after four doses of SARS-CoV-2 vaccines identified in multivariable analysis.

| Variable | Odds Ratio (95% CI) | P value |
| --- | --- | --- |
| **Low positive anti-SARS-CoV-2-S-protein IgG before vaccination** | **16.2 (3.69 - 125)** | **0.001** |
| Female sex | 1.25 (0.59 - 2.70) | 0.57 |
| **Age** | **0.96 (0.93 - 0.99)** | **0.004** |
| BMI | 1.05 (0.97 - 1.15) | 0.25 |
| mRNA vaccine | 2.19 (0.78 - 6.52) | 0.14 |
| **Transplant age** | **1.11 (1.05 - 1.18)** | **<0.001** |
| Diabetes | 1.90 (0.77 - 4.83) | 0.17 |
| CNI treatment | 0.31 (0.01 - 3.58) | 0.38 |
| Steroid treatment | 0.59 (0.07 - 4.27) | 0.60 |
| **Belatacept** | **0.01 (0.0003 - 0.17)** | **0.004** |
| MPA | 0.65 (0.07 - 5.49) | 0.69 |
| MPA dose in MMF equivalent in g | 0.31 (0.08 - 1.19) | 0.09 |
| mTORi | - | - |
| Azathioprine | - | - |
| More than 2 immunosuppressive drugs | 1.37 (0.24 - 9.12) | 0.73 |
| eGFR | 1.01 (0.99 - 1.03) | 0.23 |
| Leukocyte count | 0.95 (0.82 - 1.11) | 0.52 |
| Hemoglobin | 1.20 (0.93 - 1.57) | 0.16 |
| Albumin creatinine ratio in g/g | 0.58 (0.18 - 1.05) | 0.17 |

Belatacept treatment and higher MPA dose were associated with reduced serological response after three doses, and belatacept was associated with reduced serological response after four doses. Hence, both are the two major modifiable risk factors found in our analysis apart from repeated vaccination.

### Change in MPA dose as predictor of serological response after four doses of SARS-CoV-2 vaccine

Next, we analyzed how MPA dose adjustment affects serological response to fourth vaccination in patients receiving CNI and MPA as maintenance immunosuppression. Among 200 patients receiving a fourth dose of a SARS-CoV-2 vaccine, 33 patients maintained a steady MPA dose, 63 received a reduced MPA dose and 104 patients had paused MPA before immunization. Baseline characteristics are summarized in **Table 5** and showed higher BMI in the reduced MPA dose group in comparison to the paused MPA group (p=0.038) and higher eGFR in the reduced group in comparison to the paused and steady MPA dose groups (p=0.001, and p<0.001 respectively). Patients in the steady MPA dose group had a higher rate of low-positive IgG titers in comparison to the reduced dose group (p=0.028), and the paused MPA dose group had a higher rate of mRNA vaccinations in comparison to the other groups (each p<0.001). Mean MPA dose was 0.87 g in the reduced MPA group and 1.18 g in the steady MPA group (p<0.001).

**Table 5:** Baseline characteristics of patients in the steady MPA group, the reduced MPA group and the paused MPA group receiving a fourth dose of a SARS-CoV-2 vaccine. Continuous variables are presented as mean (+/- standard deviation) unless stated otherwise.

|  | MPA Pause | MPA Reduction | MPA Continuation | P values |
| --- | --- | --- | --- | --- |
| Number of Patients | 104 | 63 | 33 |  |
| Median age in years (IQR) | 60 (51 - 70) | 63 (47 - 70) | 64 (55 - 71) | Pause vs. cont: 0.171<br>Pause vs. red: 0.838<br>Red vs. cont: 0.265 |
| Female | 33.7% | 25.4% | 33.3% | Pause vs. cont: 0.975<br>Pause vs. red: 0.264<br>Red vs. cont: 0.417 |
| BMI in kg/m<sup>2</sup> | 25.5 +/- 4.3 | 24.2 +/- 4.3 | 24.6 +/- 4.2 | Pause vs. cont: 0.127<br>**Pause vs. red: 0.038 \***<br>Red vs. cont: 0.817 |
| Median transplant Age in years (IQR) | 7.6 (3.0 - 7.5) | 6.8 (2.1 - 11.4) | 4.9 (2.3 - 9.6) | Pause vs. cont: 0.101<br>Pause vs. red: 0.242<br>Red vs. cont: 0.563 |
| eGFR in ml/min/1.73m<sup>2</sup> | 47.6 +/- 19.0 | 55.1 +/- 17.8 | 44.7 +/- 8.1 | Pause vs. cont: 0.473<br>**Pause vs. red: 0.0099 \*\***<br>**Red vs. cont: 0.0076 \*\*** |
| Change in eGFR in ml/min/1.73m<sup>2</sup> | -1.09 +/- 6.73 | -2.47 +/- 9.30 | 0.53 +/- 5.96 | Pause vs. cont: 0.099<br>Pause vs. red: 0.4435<br>Red vs. cont: 0.1027 |
| ACR in g/g | 0.18 +/- 0.77 | 0.11 +/- 0.21 | 0.13 +/- 0.23 | Pause vs. cont: 0.938<br>Pause vs. red: 0.228<br>Red vs. cont: 0.939 |
| Change in ACR in g/g | 0.02 +/- 0.21 | 0.03 +/- 0.30 | -0.05 +/- 0.16 | Pause vs. cont: 0.358<br>Pause vs. red: 0.720<br>Red vs. cont: 0.677 |
| mRNA Vaccination | 96.2% | 76.2% | 75.8% | **Pause vs. cont: <0.001\*\*\***<br>**Pause vs. red: <0.001\*\*\*\***<br>Red vs. cont: 0.97 |
| IgG low positive before vaccination | 12.5% | 7.9% | 24.3% | Pause vs. cont: 0.105<br>Pause vs. red: 0.360<br>**Red vs. cont: 0.028 \*** |
| MPA dose in g MMF equivalent | 0 | 0.87 +/- 0.25 | 1.18 +/- 0.43 | **Pause vs. cont: <0.001 \*\*\*\***<br>**Pause vs. red: <0.001 \*\*\*\***<br>**Red vs. cont: <0.001 \*\*\*** |
| Patients with steroid treatment | 100% | 76.2% | 57.6% | **Pause vs. cont: <0.001 \*\*\*\***<br>**Pause vs. red: <0.001 \*\*\*\***<br>Red vs. cont: <0.06 |
| Steroid dose in mg methylprednisolone equivalent | 4.1 +/- 0.8 | 2.8 +/- 1.8 | 1.8 +/- 1.9 | **Pause vs. cont: <0.001 \*\*\*\***<br>**Pause vs. red: <0.001 \*\*\*\***<br>**Red vs. cont: 0.018 \*** |
| Median time since last vaccination in days (IQR) | 65 (56 - 83) | 65 (58 - 94) | 65 (48 - 87) | Pause vs. cont: 0.459<br>Pause vs. red: 0.346<br>Red vs. cont: 0.244 |

The serological response rate in the paused MPA group was 75%, which was significantly higher than in the reduced MPA group (46%, p=0.001) and the steady MPA group (52%, p=0.01) (**Figure 3**), with no significant difference between the latter two groups.

**Figure 3:**
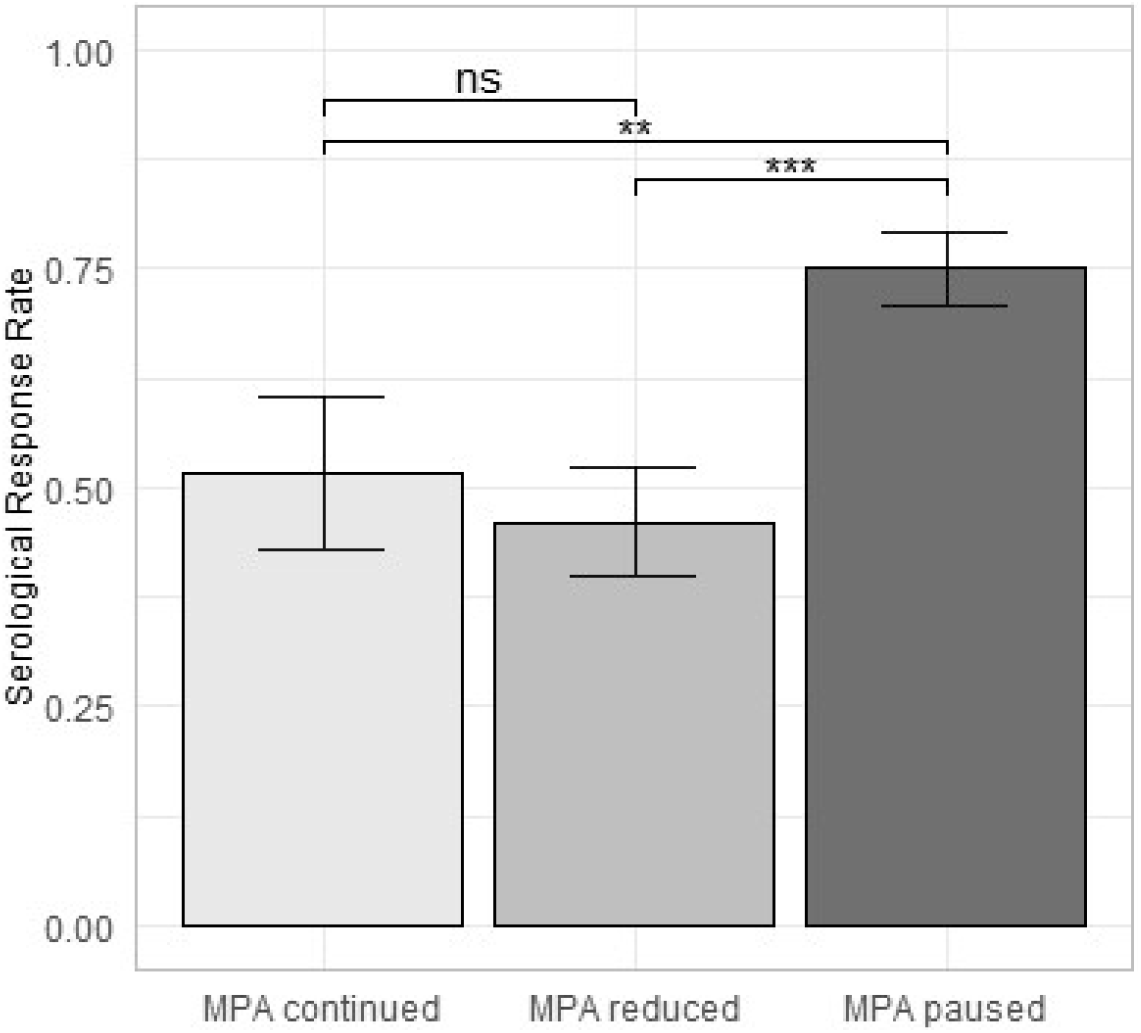
Serological response rates after four doses of SARS-CoV-2 vaccines in kidney transplant recipients with steady MPA dose (n=33), reduced MPA dose (n=63) and paused MPA (n=103).

In the paused MPA group, 1/104 patients (1%) developed de-novo DSA, and 1/104 patients (1%) developed an episode of acute T cell mediated rejection (TCMR) requiring intermittent dialysis, which could be terminated after steroid pulse therapy and adaption of immunosuppressive therapy. In the latter case, TCMR was further precipitated by two factors: first, MPA pause was extended, since the patient received abdominal wall hernia repair in another hospital, which was complicated by a superinfected hematoma; second, low tacrolimus levels of 2.59 ng/mL were found when the patient was transferred to our clinic. In the reduced MPA group, 1/63 patients (1.6%) developed de-novo DSA and 1/63 patients (1.6%) developed an episode of chronic active antibody-mediated rejection.

### Belatacept-based immunosuppression as predictor of serological response

Multivariable analysis revealed that patients, who received belatacept immunosuppression at the time of third vaccination have strongly reduced serological response. Still, we found 3 out of 63 patients (4.8%) to respond after the second vaccination, 4 out of 46 patients (8.7%) to respond after the third vaccination, 3 out of 25 patients (12%) after the fourth vaccination, and 2 out of 5 patients (40%) after the fifth vaccination. A detailed analysis revealed special immunological circumstances or reduced immunosuppressive medication in 8 out of these 9 patients with serological response, which might explain why these patients developed serological responses despite belatacept treatment (**Table S2**). Conversely, patients treated with belatacept and full dose MPA are highly unlikely to show serological response even with repeated vaccination.

In summary, patients with belatacept-based immunosuppression show impaired cumulative serological response (4.4%, 12.4%, and 16.4%) in comparison to patients with CNI-based immunosuppression (19.1%, 37.6%, and 70.1%) after basic immunization, three, and four vaccinations (**Figure 4**).

**Figure 4.**
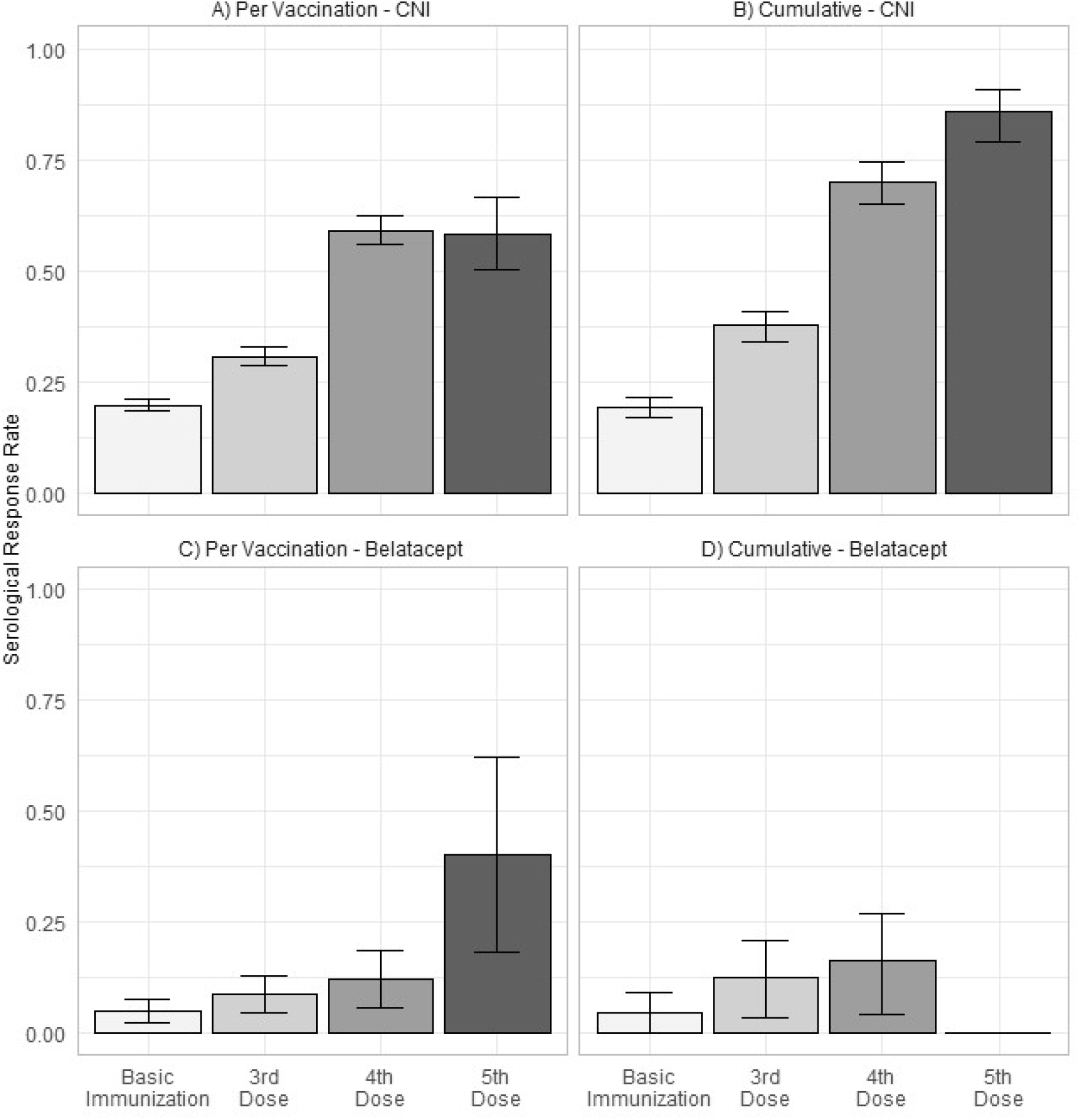
**A)** Serological response rate per vaccination (+/- standard deviation) and **B)** cumulative serological response rate (+/- 95% confidence interval) after up to 5 vaccinations in patients with CNI-based immunosuppression, as well as **C)** serological response rate per vaccination (+/- standard deviation) and **D)** cumulative serological response rate (+/- 95% confidence interval) after up to 4 vaccinations in patients with belatacept-based immunosuppression. Cumulative response rate after fifth vaccination is not shown for patients with belatacept, due to low patient count of 5 patients receiving fifth vaccination.

## Discussion

We provide the first systematic investigation analyzing the serological response to up to five repeated vaccinations against SARS-CoV-2 in a closely monitored cohort of adult KTR. It includes the largest reported cohort of KTR receiving four doses as well as the first reported cohort of KTR receiving five doses of a SARS-CoV-2 vaccine. Our data indicate that repeated vaccination of up to five times is safe and induces sufficient serological response in patients who did not respond after two or three vaccinations and achieves satisfactory antibody titers in most patients.

Contrary to other previously reported case series that supported the administration of a fourth dose of vaccine,^19,20^ we are able to also compare different approaches to the reduction of immunosuppression and their effects on serological response. In CNI-treated non-responders after three vaccinations, serological response was improved by pausing MPA and adding 5 mg prednisolone equivalent for 4 to 8 weeks at the time of fourth vaccination. A mere partial reduction of MPA, however, did not lead to an improved response rate.

In patients treated with belatacept, additional immunizations have only a limited effect on a serological response, in particular if treated with full-dose MPA – a result that complements previous descriptions of poor serological response to three doses of vaccine in KTR under belatacept immunosuppression.^21,22^ It is obvious that these patients require different approaches.

We were able to show that several factors reported to affect the response to basic immunization against SARS-CoV-2 are also predictors of serological response after three and four doses of vaccine.^23,24^ Multivariable analysis revealed that MPA dose and belatacept treatment are the most important modifiable risk factors of impaired serological response while non-modifiable factors include younger age, higher BMI and years after transplantation. These observations are consistent with the concept that increased immunosenescence in elderly individuals leads to a diminished vaccine response in general ^25-27^ and especially in KTR.^23,28^ A longer time after transplantation goes along with a general reduction of immunosuppression from waning steroids over a reduction in CNI levels.^29^ Conflicting data exist concerning the BMI of patients. On the one hand it has been shown that convalescent plasma donors with higher BMI had higher and more stable antibody titers,^30^ while a direct impact of BMI on serological response has not been constantly reported.^31,32^

We believe that this analysis has several important implications. First, the observation that patients with CNI and MPA based maintenance immunosuppression are likely to develop a serological response after four or five SARS-CoV-2 vaccines suggests that repeated vaccination is an alternative immunization strategy to the administration of monoclonal antibodies in non-responders after 3 vaccinations, with the latter being performed by several transplant centers. Second, serological response can be improved by pausing MPA and adding 5 mg prednisolone equivalent 1 week before until 4 weeks after vaccination without increased short-term risk of rejection.

Still, anti-HLA antibody development may occur with time delay in a low percentage and has to be weighed against the benefit of protection from a potentially life-threatening disease. As a consequence, we recommend close monitoring after temporary change of immunosuppression.

Third, patients receiving belatacept are likely to be sustained non-responders even after five vaccinations. For these patients, the optimal strategy to prevent severe COVID-19 has to be defined. We advocate for pre-exposure prophylaxis with monoclonal anti-S-protein antibodies for non-responders after three vaccines, who receive belatacept treatment. Switching from belatacept to CNI-based regimen is another option, but the effect of belatacept lasts two to three months after cessation. Since some patients receive belatacept as a rescue therapy in case of poor graft function (e.g. due to CNI toxicity or thrombotic microangiopathy) the optimal strategy for belatacept-treated patients remains a challenge. Other concepts might include a pill-in-the-pocket concept using nirmatrelvir/ritonavir for early treatment after exposure. This needs to be performed under dose adaption or pause of CNI and close monitoring of CNI levels. Alternatively, early treatment with remdesevir or post-exposure prophylaxis with monoclonal antibodies are more widely available alternatives.

Limitations arise from the study’s retrospective design. While serological measurements were routinely performed at our institution, there was a considerable number of vaccinations that were not preceded or followed by serological measurements (**Figure 1**). Consequently, a patient-based approach was discarded in favor of a vaccination-based evaluation, introducing a risk of selection bias. However, our approach allows to account for the incompleteness of the data and to optimize the number of examinable vaccinations. Intraindividual changes of titers between or after vaccinations were not represented with this approach, whereas protection after vaccination will evolve and might effectively decrease over time.

With regard to the effects of MPA dose adjustment around the fourth vaccination, the retrospective assignment to treatment groups limits the validity of the results because groups were not fully matched and confounding factors could have influenced the group assignment. Nevertheless, comparison of the major potential influencing factors was provided to account for these risks.

Finally, while this study focuses on vaccine-induced humoral response as correlate of protection from disease, there are other contributors to immunity such as T cell response that may influence the degree of protection.

In conclusion, repeated vaccination against SARS-CoV-2 of up to five times effectively induces humoral serological response in kidney transplant recipients. Serological response can be enhanced by pausing MPA at the time of vaccination without increased short-term risk of acute rejection. Patients with belatacept immunosuppression and full-dose MPA are unlikely to achieve sufficient serological response, thus requiring a different approach to ensure protection for this population at-risk.

## Data Availability

All data produced in the present study are available upon reasonable request to the authors.

## Abbreviations

CNI: calcineurin inhibitor
COVID-19: coronavirus disease 2019
eGFR: estimated glomerular filtration rate
ELISA: enzyme-linked immunosorbent assays
KTR: kidney transplant recipients
MPA: mycophenolic acid
N: nucleocapsid protein
RBD: receptor binding domain
S: spike protein
SARS-CoV-2: severe acute respiratory syndrome coronavirus 2
SOT: solid organ transplantation
TCMR: T cell mediated rejection

## Authors’ Contributions

ESc, BO, SR, and KB conceived of the presented idea. BO and SR performed data analysis. FG and MM assisted in data analysis. SR, BO, and ESc wrote the manuscript. AJ, ESc, KB, FH, FB, MC, UW, BZ and CH performed vaccinations. NK, ESe, BZ, JW, FB, FH, UW, MC, and KB performed follow-up visits. KB, LL, FH, and KUE provided significant intellectual input during the conception and development of the article. BE and JH are responsible for the serological testing. All authors commented and reviewed the final manuscript.

## Acknowledgements

ESc is participant in the BIH-Charité Clinician Scientist Program funded by the Charité– Universitätsmedizin Berlin and the Berlin Institute of Health.

## Disclosures

Nothing to disclose.

## Funding

No funding.

## Data Sharing Statement

All data produced in the present study are available upon reasonable request to the authors.

## Supplemental Table of Contents

**Item S1:** Detailed description of institutional SARS-CoV-2 vaccination protocol.

**Figure S1:** Graphical representation of the institutional SARS-CoV-2 vaccination protocol.

**Table S1:** Variable definition for descriptive statistics and multivariable analysis.

**Table S2**: Detailed analysis of all belatacept patients with sufficient serological response showing special immunological circumstances or reduced immunosuppressive medication in 8 out of 9 patients.

## Notes

### Competing Interest Statement

The authors have declared no competing interest.

### Funding Statement

This study did not receive any funding.

### Author Declarations

The ethics committee of Charité - Universitätsmedizin Berlin gave ethical approval for this work (EA1/030/22)

