## Supplementary Data for "Serological Response to Three, Four and Five Doses of SARS-CoV-2 Vaccine in Kidney Transplant Recipients"

**Item S1:** Detailed description of institutional SARS-CoV-2 vaccination protocol.

**Figure S1:** Graphical representation of the institutional SARS-CoV-2 vaccination protocol.

**Table S1:** Variable definition for descriptive statistics and multivariable analysis.

**Table S2**: Detailed analysis of all belatacept patients with sufficient serological response showing special immunological circumstances or reduced immunosuppressive medication in 8 out of 9 patients.

**Item S1:**

As per institution protocol, patients, who had no sufficient serological response 4-8 weeks after basic immunization, received a third vaccination dose. Patients who did not show sufficient serological response 4-8 weeks after third vaccination dose and received CNI + MPA immunosuppression, were treated either with continuation of MPA dose, reduction of MPA dose of 25-50%, or MPA pause with additional 5 mg prednisolone equivalent from 4-7 days before, and until 2-4 weeks after the next vaccination. The individual regimen was dependent on patient-related factors (such as previous MPA dose, erythrocyte IMPDH activity [1], presence of DSA, triple vs. dual immunosuppression, previous rejection episodes, and patient preference) as well as the treating physician. While some physicians performed no MPA pauses, others performed them regularly for fourth and fifth vaccination.

For patients receiving belatacept maintenance immunosuppression without sufficient serological response, a fourth and fifth vaccination dose was administered without a change in immunosuppression before December 2021, since most patients were receiving Belatacept as “rescue therapy” [2-4] and at that time no monoclonal antibodies were available for pre-exposure prophylaxis at our institution.

After December 2021, patients with belatacept immunosuppression without sufficient serological response after third, fourth or fifth vaccination dose received pre-exposure prophylaxis with monoclonal antibodies. This explains, why only few patients with belatacept immunosuppression received fifth vaccination without meeting the exclusion criteria (one of which was treatment with monoclonal antibodies before vaccination response).

**Figure S1:**

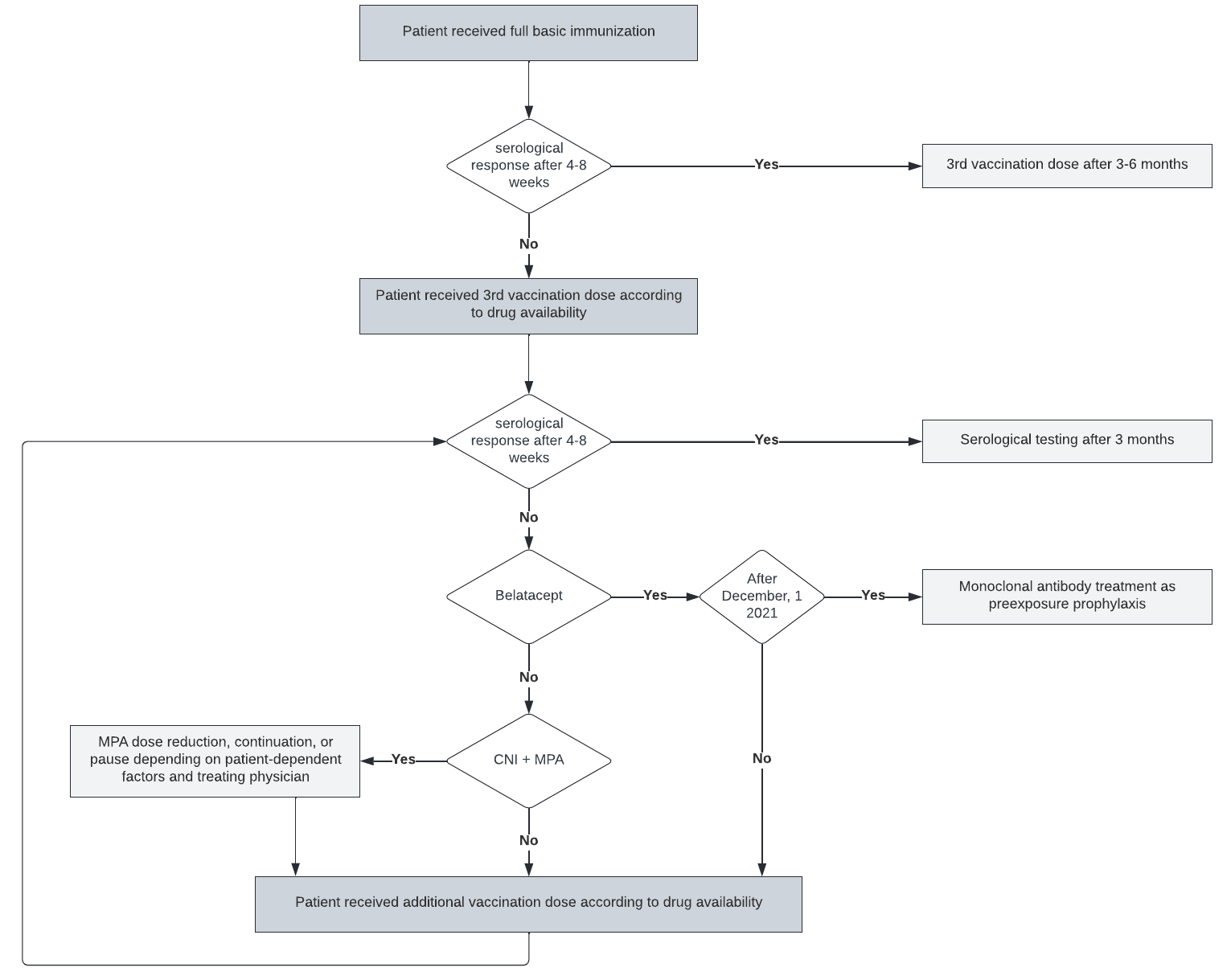

**Table S1:**

| **Candidate Variable** | **Data Type** | **Definition** |
| --- | --- | --- |
| Sex | binary (f/m) | sex (female or male) |
| Age | float | age at the time of vaccination in years |
| BMI | float | body mass index (body mass divided by square of the body height) in kg/m^2^ |
| mRNA vaccine | binary (1/0) | respective vaccination performed with an mRNA-based SARS-CoV-2 vaccine |
| Transplant age | float | time since the patient's last kidney transplantation at the time of vaccination in years |
| Diabetes | binary (1/0) | a diagnosis of diabetes mellitus in the patient's history or current use of antidiabetic medication |
| CNI treatment | binary (1/0) | use of systemic tacrolimus or cyclosporine as an immunosuppressive medication at the time of vaccination |
| Steroid treatment | binary (1/0) | use of systemic steroids as an immunosuppressive medication at the time of vaccination |
| Belatacept | binary (1/0) | use of belatacept as an immunosuppressive medication at the time of vaccination |
| MPA | binary (1/0) | use of mycophenolic acid (as mycophenolate sodium or mycophenolate mofetil (MMF)) as an immunosuppressive medication at the time of vaccination |
| MPA dose | float | mycophenolic acid dose in MMF equivalent in g at the time of vaccination |
| mTORi | binary (1/0) | use of systemic sirolimus or everolimus as an immunosuppressive medication at the time of vaccination |
| Azathioprine | binary (1/0) | use of systemic azathioprine as an immunosuppressive medication at the time of vaccination |
| More than 2 immunosuppressive drugs | binary (1/0) | use of more than two types of immunosuppressive medication at the time of vaccination |
| eGFR | float | most recent estimated glomerular filtration rate according to the Chronic Kidney Disease Epidemiology Collaboration (CKD-EPI) equation in ml/min/1.73m² at the time of vaccination (if unavailable, first measurement after vaccination) |
| Leukocytes | float | most recent leukocyte count in /nl at the time of vaccination (if unavailable, first measurement after vaccination) |
| Hemoglobin | float | most recent hemoglobin level in mg/dL at the time of vaccination (if unavailable, first measurement after vaccination) |
| Urine albumin creatinine ratio | float | most recent urinary albumin-to-creatinine ratio (ACR) in either spot or  24 hour collection urine in g/g at the time of vaccination (if unavailable, first measurement after vaccination) |

**Table S2**:

| Patient No. | Vaccination No. | Potential explanation for improved vaccine response |
| --- | --- | --- |
| 1 | 3 | Vaccination at time of belatacept uptitration, no MPA due to severe diarrhea |
| 2 | 3 | Belatacept and steroid only, no MPA |
| 3 | 3 | Patient received basic immunization before kidney transplantation, but returned negative after transplantation and switch to belatacept. |
| 4 | 4 | Low MPA dose (500mg/d MMF equivalent) |
| 5 | 4 | Low MPA dose (500mg/d MMF equivalent) and reduced dose of belatacept (4 mg/kg per 4 weeks) due to recurrent infections |
| 6 | 4 | Low MPA dose (500mg/d MMF equivalent) |
| 7 | 5 | Low MPA dose (500mg/d MMF equivalent) |
| 8 | 5 | No MPA due to BK virus infection |
| 9 | 3 | No specific explanation, standard immunosuppression. |
